# Prevalence and predictors of HIV testing among young men in Papua New Guinea: a cross-sectional analysis of a nationally representative sample

**DOI:** 10.1101/2023.03.17.23287339

**Authors:** McKenzie Maviso

## Abstract

**Background:** HIV testing is an important component of HIV prevention and serves as a gateway to other HIV-related services. However, the uptake remains suboptimal among young people, particularly in highly prevalent settings such as Papua New Guinea (PNG). This study used nationally representative data to assess the prevalence and determine predictors of HIV testing uptake among young men aged 15–24 years in PNG.

**Methods:** The 2016–2018 PNG Demographic and Health Survey (PNGDHS) data was used. A total of 1,275 young men aged 15–24 years were included in the final analysis. Descriptive, bivariate, and multivariable logistic regression analyses were performed to determine independent predictors of HIV testing. Adjusted odds ratios (AORs) with 95% confidence intervals (CIs) were reported. All analyses were adjusted using survey weights to account for unequal sampling probabilities.

**Results:** The overall prevalence of HIV testing among young men was 17.2% (95% CI: 15–19). Of those who were tested for HIV, most lived in rural areas (73%), about one-third had a sexual debut at age <15 years (32.9%), and 33.9% inconsistently used condoms during sex. In multivariable analysis, men aged 20–24 years (AOR 1.18, 95% CI: 1.00–2.31), who own mobile phones (AOR 1.43, 95% CI: 1.00– 2.55), who were aware that consistent condom use during sex can reduce HIV risk (AOR 2.18, 95% CI: 1.18–4.04), who had paid for sex (AOR 1.75, 95% CI: 1.01–5.83), and who had two or more sexual partners (AOR 1.37, 95% CI: 1.01–3.14) had increased odds of HIV testing. However, decreased odds of HIV testing were found among men who were never married (AOR 0.51, 95% CI: 0.29–0.88), lived in rural areas (AOR 0.54, 95% CI: 0.32–0.92), and consistently used condoms during sex (AOR 0.59, 95% CI: 0.34–1.01).

**Conclusion:** The findings show that HIV testing is low among young men in PNG. To increase HIV testing uptake among young men, it is crucial to implement comprehensive youth-friendly HIV/STI education and tailored sensitization programs and enable more accessible and affordable HIV testing services. Also, outreach and community-based testing programs for young men in prioritized areas requiring urgent prevention intervention are feasible options in PNG.

## Introduction

Considerable progress has been made in responding to HIV/AIDS under the Sustainable Development Goals framework to end the epidemic by 2030 [1]. Since 2010, new HIV infection rates have decreased by 14%, and 78% of people living with HIV know their status [2]. However, in the Asia and Pacific region, the HIV response is impacted by marked disparities and varied epidemic trends, notably affecting young people aged 15–24 years, who represent a growing proportion of people who disproportionately endure the burden of HIV. In 2022, an estimated 440,000 young people were living with HIV, accounting for about 25% of new HIV infections in the region [2].

The United Nations’ 95-95-95 targets to end the HIV epidemic by 2030 called for 95% of people living with HIV to know about their serostatus [3,4]. HIV testing is a key strategic entry point to prevention, treatment, care, and support services. It confers multiple benefits for individuals who test positive and those who test negative and encourages preventive behaviors [5,6]. Despite the significant benefits, young people remain the least likely to get an HIV test and know their HIV status [15,27]. Studies have shown that not all, particularly men willingly seek HIV testing services as a way to prevent HIV transmission and/or acquisition due to various barriers, such as individual (poor HIV/AIDS knowledge, testing availability, and unwillingness to test), social (stigma, discrimination, and lack of social support), and health service utilization (loss of trust or confidence in health workers and lack of medication, including ART) [7–9]. These barriers undermine efforts to increase HIV testing and highlight the critical need to develop appropriate HIV prevention strategies.

Among young people, their sexual behaviors and susceptibility to HIV infection differ widely by social context, as well as increased individual autonomy and a lack of social control [10,11]. For example, high-risk behaviors such as early sexual debut, substance use (alcohol consumption and drug use), inconsistent condom use, and multiple and concurrent sexual relationships, as well as poor HIV knowledge, low-risk perception, and sensation-seeking behaviors, increase HIV risks and transmissions among young people [12–15]. In addition, biological factors such as low rates of male circumcision [16,17] and manifestations of other sexually transmitted infections (STIs) (chlamydia, gonorrhea, syphilis, and trichomoniasis) [18,19] contribute to some of the increased HIV risk and burden among this population. Despite young people’s vulnerability to HIV infection, they are frequently overlooked in the design and implementation of national AIDS strategies [20,21]. They remain an important target group for HIV prevention and STI surveillance due to their unique behavioral and social-related vulnerability.

PNG is one of the culturally diverse countries in the Western Pacific region and is grappling with a concentrated-generalized HIV epidemic, indicating significant disparities in HIV prevalence between key populations (e.g., sex workers and men who have sex with men) and the general population [22,23]. HIV prevalence is 1% in the general adult population [24]. Among young people, the prevalence of HIV is estimated at 0.2% for males and 0.5% for females [24], demonstrating the high vulnerability of this priority group in the country. Although HIV testing services are widely available throughout PNG [25], the current testing rate among young people is suboptimal, especially among those most at risk of HIV [26,27]. Furthermore, a major obstacle in the country continues to be young men’s participation in HIV programs and their access to sexual and reproductive health care [28,29]. Evidence concerning HIV knowledge and risk reduction behaviors (e.g., consistent use of condoms and having only one faithful sexual partner) among young men is scarce; although a recent study among young women in PNG found that early sexual debut, having one sexual partner and not having an STI were associated with not testing for HIV [26]. However, young men’s knowledge of HIV and sexual relationships is established through a complex interplay of sociocultural norms, expectations, attitudes, and information gleaned from various sources [30].

Despite the established profile of risky sexual behaviors among young people [26,31,32], little is known about the prevalence and correlates of HIV testing uptake in young men in PNG. Moreover, evidence concerning predictors influencing the uptake of HIV testing among young men is scarce. This creates a critical knowledge gap that must be filled to inform national-level policy development and targeted interventions. In this study, data from a nationally representative survey was used to determine the prevalence and predictors of HIV testing uptake among young men aged 15–24 years in PNG.

## Methods and materials

### Data sources

Data for the current study were derived from the 2016–2018 PNG Demographic and Health Survey (PNGDHS), which used a stratified, multistage cluster sampling method to ensure a sample representative of the population. The survey was conducted between October 2016 and December 2018 and collected data on the country’s important demographic, socioeconomic, and health indicators. The survey used the list of census units (CUs) from the 2011 PNG National Population and Housing Census [33] as the sampling frame, which contains information on CU location, type of residence (urban or rural), estimated number of residential households, and population by sex. Administratively, the country is divided into four (4) main regions (Southern, Highlands, Momase, and Islands) comprising 22 provinces, and each province is subdivided into urban and rural areas. The PNG National Statistical Office (NSO) with technical support and assistance from the Inner City Fund (ICF) through the DHS Program conducted the study. The governments of PNG and Australia, the United Nations Population Fund (UNFPA), and the Children’s Fund of the United Nations (UNICEF) funded the survey [34].

### Sampling procedures

The 2016–2018 PNGDHS sample stratification and selection were achieved in two strata: urban and rural areas, except for the National Capital District, which has no rural areas. In each stratum, samples of households were selected in two stages, with CU constituting the primary sampling unit and households as the secondary sampling unit. The first stage involved selecting 800 CUs using a probability proportional to the CU size. In the second stage, 24 households were selected from each cluster using probability sampling, yielding a total sample size of approximately 19,200 households. A total of 7,333 men aged 15–49 years participated in the survey. Young men who never had sex and aged 25 or more were excluded from this study. A total weighted sample of 1,275 young men aged 15– 24 years who ever had sex with variables of interest were included in this study. Details about the sampling technique, survey team training, household selection, survey questionnaires, and validation procedures are available in the final report [34].

### Variables and measurements

The primary outcome variable of this study was “ever tested for HIV?” and was dichotomized as “0” for “no” or “1” for “yes.” The explanatory variables were selected based on the literature and their relevance [18–20] and were divided into three categories: (1) Sociodemographic factors include age, marital status, educational level, employment status, wealth index, region, and place of residence. Also, household factors measured included exposure to mass media (listening to the radio and watching television), access to the internet, and mobile phone ownership. (2) HIV-related knowledge includes ever heard of AIDS or STIs, can a healthy person have HIV? Can a person get HIV by sharing food with an infected person? Can a person get HIV through witchcraft? Consistent condom use can reduce HIV risk and knowing an HIV testing facility. (3) Sexual risk behaviors include age of sexual debut, number of sexual partners (including spouse), having ever paid for sex, and consistent condom use during sex, and having had an STI (unspecific) in the last 12 months.

### Statistical analysis

Data extraction, recoding, and analysis were performed using IBM Statistical Package for the Social Sciences (SPSS), Version 26.0 (Armonk, NY: IBM Corp.). The sample was weighted using the primary sampling unit variable, stratification variable, and weight variable to restore its representativeness and obtain a better estimate throughout the analysis. Descriptive statistics were used to explore the characteristics of the sample and were presented as weighted frequencies (*n*) and percentages (%). A chi-square test for independence was performed to assess the strength of the association between the dependent variable and each independent variable. Variables that were significant and those with *p* ≤ 0.25 in the bivariate analysis were included in a multivariable analysis to determine their collective associations with HIV testing [38]. Since the PNGDHS used a two-stage stratified sampling technique, a complex samples analysis technique was employed to determine independent predictors of HIV testing. Adjusted odds ratios (AORs) with 95% confidence intervals (CIs) were reported. A *p* ≤ 0.05 was used to assess statistical significance.

### Ethical Considerations

Permission to use data was obtained from the DHS program (https://dhsprogram.com) and was only used for this study. The Institutional Review Board (IRB) of Inner City Fund (ICF) International and the IRB committee of PNG examined and approved the 2016–2018 PNGDHS protocols. All the respondents had provided verbal informed consent before each interview. There were no ethical considerations on the researcher’s part as the data were anonymized entirely, with no identifiable information on the survey participants when analyzed.

## Results

### Sociodemographic characteristics of young men

Table 1 presents the participants’ characteristics. Overall, 1,275 sexually active young men aged 15–24 years enrolled in the study. The mean age was 20.84 (± 2.31) years, while the majority (70.2%) were aged 20–24 years and lived in rural areas (82%). Nearly half had attained primary (44.3%) and secondary (45.5%) education, and about two-thirds (62%) were unemployed. Regarding mass media exposure, more than two-thirds of young men listened to the radio (71.7%), while less than half watched television (47.1%) and had internet access (33.6%). More than half (57.8%) of young men owned mobile phones.

**Table 1.** Sociodemographic characteristics of participants (N = 1,275)

| Characteristics | Frequency |  |
| --- | --- | --- |
|  | *n* | % |
| *Sociodemographic* |  |  |
| Age (years) |  |  |
| 15–19 | 380 | 29.8 |
| 20–24 | 895 | 70.2 |
| Marital status (missing, *n* = 26) |  |  |
| Never married | 940 | 75.2 |
| Married | 310 | 24.8 |
| Educational level |  |  |
| No formal education | 73 | 5.7 |
| Primary | 565 | 44.3 |
| Secondary | 580 | 45.5 |
| Tertiary | 58 | 4.5 |
| Employment status |  |  |
| Not employed | 791 | 62.0 |
| Employed | 485 | 38.0 |
| Wealth Index |  |  |
| Poorest | 120 | 9.4 |
| Poorer | 119 | 9.3 |
| Middle | 189 | 14.8 |
| Richer | 294 | 23.0 |
| Richest | 553 | 43.4 |
| Place of residence |  |  |
| Urban | 229 | 18.0 |
| Rural | 1,046 | 82.0 |
| Region |  |  |
| Southern | 300 | 23.6 |
| Highlands | 455 | 35.7 |
| Momase | 373 | 29.2 |
| Islands | 147 | 11.5 |
| Read newspaper/magazine |  |  |
| No | 369 | 28.9 |
| Yes | 907 | 71.1 |
| Listen to the radio |  |  |
| No | 361 | 28.3 |
| Yes | 914 | 71.7 |
| Watch television |  |  |
| No | 674 | 52.9 |
| Yes | 601 | 47.1 |
| Internet access |  |  |
| No | 847 | 66.4 |
| Yes | 428 | 33.6 |
| Own a mobile phone |  |  |
| No | 537 | 42.2 |
| Yes | 738 | 57.8 |

**Note:** Weighted frequencies (*n*) and percentages (%)

### The prevalence of having ever been tested for HIV

Overall, 17.1% (95% CI: 0.15, 0.19) of the young men had ever been tested for HIV (Fig 1).

**Fig 1.**
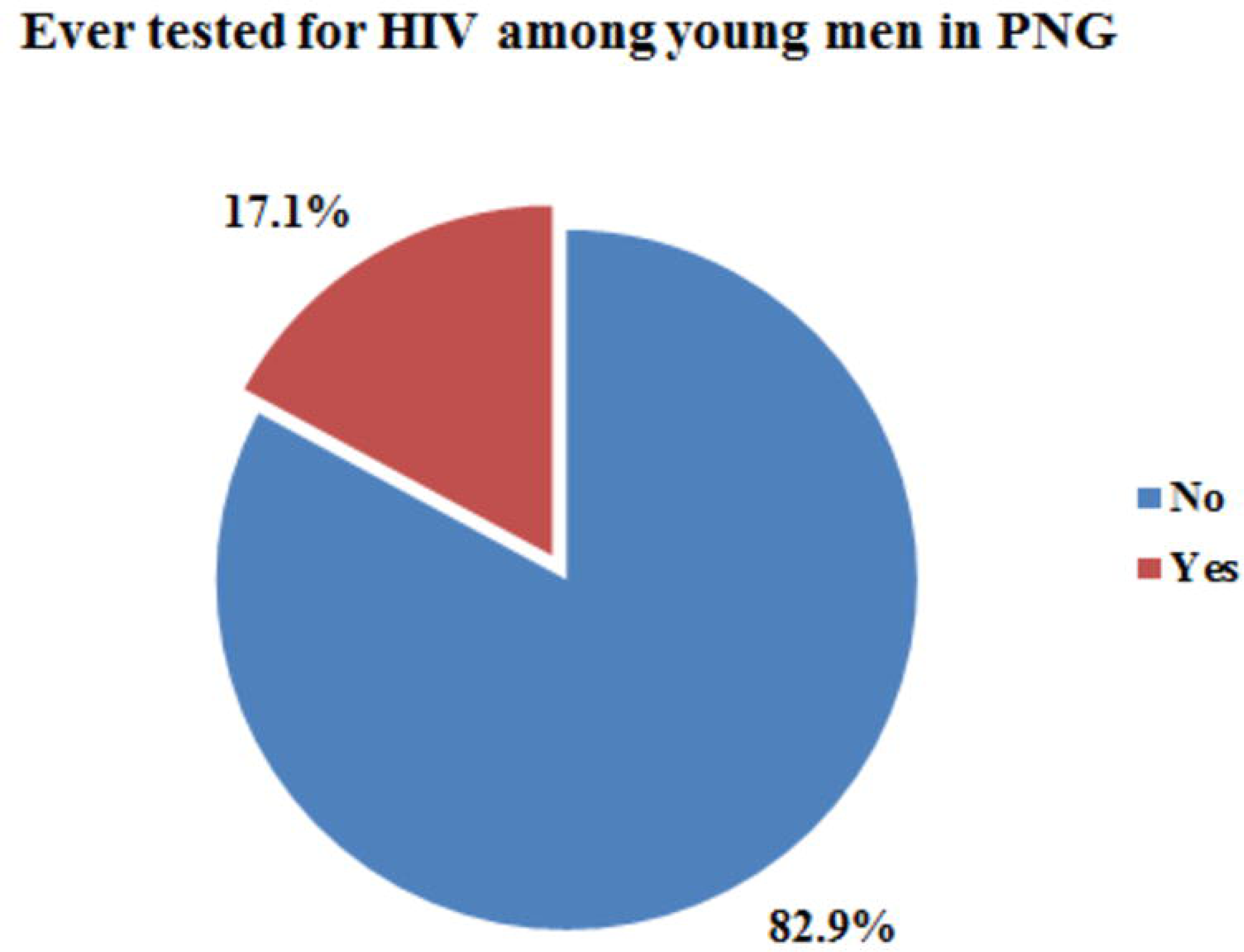
The rates of HIV testing among young men in PNG.

### HIV knowledge and sexual risk behaviors

Table 2 shows the HIV-related knowledge and sexual risk behaviors of young men. The vast majority of young men had adequate knowledge that a healthy person can have HIV (90.8%) and that using consistent condom use during sex can reduce the risk of HIV transmission (80.2%). Most knew of an HIV testing facility (70.8%). Regarding young men’s sexual risk behaviors, over a quarter were reported to have initiated sex at an early age (<15 years) (28.2%) and inconsistently used condoms during sex (27.7%). The majority of young men had a single sexual partner in the last 12 months (88.7%).

**Table 2.** HIV knowledge and sexual risk behaviors of young men (N = 1,275)

| Characteristics | Frequency |  |
| --- | --- | --- |
|  | *n* | % |
| ***HIV Knowledge*** |  |  |
| Ever heard of AIDS |  |  |
| No | 92 | 7.2 |
| Yes | 1,183 | 92.8 |
| Ever heard of STIs |  |  |
| No | 74 | 5.8 |
| Yes | 1,201 | 94.2 |
| A healthy person can have HIV (missing, *n* = 326) |  |  |
| No | 88 | 9.2 |
| Yes | 861 | 90.8 |
| A person can get HIV by sharing food with an infected person (missing, *n* = 298) |  |  |
| No | 822 | 84.1 |
| Yes | 155 | 15.9 |
| A person can get HIV through witchcraft (missing, *n* = 297) |  |  |
| No | 908 | 92.8 |
| Yes | 70 | 7.2 |
| Consistent condom use during sex can reduce HIV risk (missing, *n* = 311) |  |  |
| No | 191 | 19.8 |
| Yes | 773 | 80.2 |
| Know HIV testing facility (missing, *n* = 93) |  |  |
| No | 346 | 29.2 |
| Yes | 836 | 70.8 |
| ***Sexual Risk Behavior*** |  |  |
| Age of sexual debut (years) |  |  |
| <15 | 360 | 28.2 |
| >15 | 916 | 71.8 |
| Number of sexual partners, including spouse (last 12 months) |  |  |
| 0–1 | 1,130 | 88.7 |
| 2 or more | 144 | 11.3 |
| Ever paid for sex (missing, *n* = 49) |  |  |
| No | 1,164 | 94.9 |
| Yes | 63 | 5.1 |
| Consistent condom use during sex (missing, $n = 95$) | | |
| No | 326 | 27.7 |
| Yes | 854 | 72.3 |
| Had any STIs (last 12 months)<sup>†</sup> |  |  |
| No | 1,211 | 94.9 |
| Yes | 65 | 5.1 |

The type of STI was not specified in the dataset.
**Note:** Weighted frequencies ( $n$ ) and percentages (%)

### Bivariate analysis of predictors of HIV testing among young men

Table 2 presents the relationship between sociodemographic characteristics, HIV knowledge, sexual risk behaviors, and HIV testing uptake among young men. Overall, only 17.1% (*n* = 222) of young men have ever been tested for HIV across sociodemographic, HIV knowledge, and sexual risk behavior strata. Having ever been tested for HIV was high among men aged 20–24 years (78.8%), those who were married (66.5%), had attained secondary education (58.1%), were unemployed (53.6%), and lived in rural areas (73%). Similarly, HIV testing remained high among those who read a newspaper or magazine (81.5%), listened to the radio (79.2%), and watched television (53.2%). Regarding HIV knowledge, having ever heard of AIDS and STIs (*p*<0.001), being aware that consistent use of condoms can reduce HIV risk (*p* = 0.011), and knowing an HIV testing facility (*p*<0.001) were significantly associated with HIV testing uptake. Furthermore, there was a statistically significant association between HIV testing and the following sexual risk behavior factors: age of sexual debut (*p* = 0.049), ever paid for sex (*p*<0.001), consistent condom use (*p* = 0.02), and having an STI in the past 12 months (*p*<0.001).

### Multivariable analysis of predictors of HIV testing among young men

Table 3 shows the results of the multivariable analysis examining the association between HIV testing uptake and sociodemographic, HIV-related knowledge, and sexual risk behavior variables. Age, marital status, place of residence, being aware that consistent condom use during sex can reduce HIV risk, ever paid for sex, number of sexual partners, and consistent condom use during sex were statistically significant predictors of HIV testing uptake among young men. In the multivariable analysis, men aged 20–24 years (AOR 1.18, 95% CI: 1.00–2.31), who own mobile phones (AOR 1.43, 95% CI: 1.00– 2.55), who were aware that consistent condom use during sex can reduce HIV risk (AOR 2.18, 95% CI: 1.18–4.04), who had paid for sex (AOR 1.75, 95% CI: 1.01–5.83), and who had two or more sexual partners (AOR 1.37, 95% CI: 1.01–3.14) had increased odds of HIV testing. However, decreased odds of HIV testing uptake were found among men who were never married (AOR 0.51, 95% CI: 0.29– 0.88), lived in rural areas (AOR 0.54, 95% CI: 0.32–0.92), and consistently used condoms during sex (AOR 0.59, 95% CI: 0.34–1.01).

**Table 3.** Bivariate analysis of sociodemographic, HIV knowledge, behavior, and HIV testing (N = 1,275)

| Characteristics | Ever been tested for HIV |  |  |  | P-value |
| --- | --- | --- | --- | --- | --- |
|  | No<br>(n = 1,053) | % | Yes<br>(n = 222) | % |  |
| *Sociodemographics* |  |  |  |  |  |
| Age (years) |  |  |  |  | 0.002* |
| 15–19 | 333 | 31.6 | 47 | 21.1 |  |
| 20–24 | 720 | 68.4 | 175 | 78.8 |  |
| Marital status (missing, n = 26) |  |  |  |  | 0.001** |
| Never married | 238 | 23.0 | 72 | 33.5 |  |
| Married | 796 | 77.0 | 143 | 66.5 |  |
| Educational level |  |  |  |  | 0.000** |
| No formal education | 68 | 6.5 | 5 | 2.3 |  |
| Primary | 490 | 46.5 | 75 | 33.8 |  |
| Secondary | 450 | 42.7 | 129 | 58.1 |  |
| Tertiary | 45 | 4.3 | 13 | 5.9 |  |
| Employment status |  |  |  |  | 0.005* |
| Not employed | 671 | 63.7 | 119 | 53.6 |  |
| Employed | 382 | 36.3 | 103 | 46.4 |  |
| Wealth Index |  |  |  |  | 0.271 |
| Poorest | 96 | 9.1 | 24 | 10.9 |  |
| Poorer | 94 | 8.9 | 25 | 11.3 |  |
| Middle | 162 | 15.4 | 27 | 12.2 |  |
| Richer | 235 | 22.3 | 58 | 26.2 |  |
| Richest | 466 | 44.3 | 87 | 39.4 |  |
| Place of residence |  |  |  |  | 0.000** |
| Urban | 169 | 16.1 | 60 | 27.0 |  |
| Rural | 883 | 83.9 | 162 | 73.0 |  |
| Region |  |  |  |  | 0.000** |
| Southern | 248 | 23.6 | 53 | 23.8 |  |
| Highlands | 355 | 33.7 | 100 | 44.8 |  |
| Momase | 332 | 31.5 | 41 | 18.4 |  |
| Islands | 118 | 11.2 | 29 | 13.0 |  |
| Read newspaper/magazine |  |  |  |  | 0.000** |
| No | 328 | 31.1 | 41 | 18.5 |  |
| Yes | 725 | 68.9 | 181 | 81.5 |  |
| Listen to the radio |  |  |  |  | 0.006* |
| No | 315 | 29.9 | 46 | 20.8 |  |
| Yes | 738 | 70.1 | 175 | 79.2 |  |
| Watch television |  |  |  |  | 0.048* |
| No | 570 | 54.1 | 104 | 46.8 |  |
| Yes | 483 | 45.9 | 118 | 53.2 |  |
| Internet access |  |  |  |  | 0.000** |
| No | 730 | 69.3 | 117 | 52.7 |  |
| Yes | 323 | 30.7 | 105 | 47.3 |  |
| Own a mobile phone |  |  |  |  | 0.000** |
| No | 475 | 45.1 | 62 | 28.1 |  |
| Yes | 578 | 54.9 | 159 | 71.9 |  |
| ***HIV Knowledge*** |  |  |  |  |  |
| Ever heard of AIDS |  |  |  |  | 0.000** |
| No | 92 | 8.7 | 0 | 0 |  |
| Yes | 961 | 91.3 | 222 | 100 |  |
| Ever heard of STIs |  |  |  |  | 0.000** |
| No | 47 | 7.0 | 0 | 0 |  |
| Yes | 979 | 93.0 | 222 | 100 |  |
| A healthy person can have HIV (missing, $n = 326$) | | | | | 0.170 |
| No | 77 | 9.9 | 11 | 6.5 |  |
| Yes | 702 | 90.1 | 158 | 93.5 |  |
| A person can get HIV by sharing food with an infected person (missing, $n = 298$) | | | | | 0.248 |
| No | 673 | 83.5 | 148 | 87.1 |  |
| Yes | 133 | 16.5 | 22 | 12.9 |  |
| A person can get HIV through witchcraft (missing, $n = 297$) | | | | | 0.376 |
| No | 739 | 92.5 | 168 | 94.4 |  |
| Yes | 60 | 7.5 | 10 | 5.6 |  |
| Consistent condom use during sex can reduce HIV risk (missing, $n = 311$) | | | | | 0.011* |
| No | 170 | 21.3 | 21 | 12.7 |  |
| Yes | 627 | 78.7 | 145 | 87.3 |  |
| Know HIV testing facility (missing, $n = 93$) | | | | | |
| No | 346 | 36.0 | 0 | 0 | 0.000** |
| Yes | 614 | 64.0 | 222 | 100 |  |
| ***Sexual Risk Behaviors*** |  |  |  |  |  |
| Age of sexual debut (years) |  |  |  |  | 0.049* |
| <15 | 287 | 27.3 | 73 | 32.9 |  |
| >15 | 766 | 72.7 | 149 | 67.1 |  |
| Number of sexual partners, including spouse (in the last 12 months) |  |  |  |  | 0.101 |
| 0–1 | 941 | 89.4 | 189 | 85.5 |  |
| 2 or more | 112 | 10.6 | 32 | 14.5 |  |
| Ever paid for sex (missing, *n* = 49) |  |  |  |  | 0.000** |
| No | 971 | 96.7 | 192 | 88.1 |  |
| Yes | 37 | 3.7 | 26 | 11.9 |  |
| Consistent condom use during sex (missing, *n* = 95) |  |  |  |  | 0.020* |
| No | 251 | 26.2 | 75 | 33.9 |  |
| Yes | 707 | 73.8 | 146 | 66.1 |  |
| Had any STIs (last 12 months)<sup>†</sup> |  |  |  |  | 0.000** |
| No | 1,010 | 95.9 | 200 | 90.1 |  |
| Yes | 43 | 4.1 | 22 | 9.9 |  |

\* $p \leq 0.05$
\*\* $p \leq 0.001$
<sup>†</sup>The type of STI was not specified in the dataset.

**Table 3.** Multivariable analysis of predictors of HIV testing among young men (N = 1,275)

| Characteristics | Ever been tested for HIV |  |  |  | *p*-value |
| --- | --- | --- | --- | --- | --- |
|  | COR | 95% CI | AOR | 95% CI |  |
| Age (years) |  |  |  |  | 0.026* |
| 15–19 | Ref |  | Ref |  |  |
| 20–24 | 1.73 | 1.09–2.76 | 1.18 | 1.00–2.31 |  |
| Marital status |  |  |  |  | 0.016* |
| Never married | 0.59 | 0.39–0.89 | 0.51 | 0.29–0.88 |  |
| Married | Ref |  | Ref |  |  |
| Educational level |  |  |  |  | 0.749 |
| No formal education | 0.25 | 0.05–1.09 | 0.83 | 0.13–5.33 |  |
| Primary | 0.53 | 0.18–1.58 | 1.23 | 0.34–4.49 |  |
| Secondary | 0.98 | 0.31–3.09 | 1.55 | 0.42–5.69 |  |
| Tertiary | Ref |  | Ref |  |  |
| Employment status |  |  |  |  | 0.380 |
| Not employed | 0.66 | 0.45–0.97 | 0.78 | 0.45–1.36 |  |
| Employed | Ref |  | Ref |  |  |
| Place of residence |  |  |  |  | 0.023* |
| Urban | Ref |  | Ref |  |  |
| Rural | 0.52 | 0.35–0.77 | 0.54 | 0.32–0.92 |  |
| Region |  |  |  |  | 0.313 |
| Southern | 0.87 | 0.52–1.46 | 1.19 | 1.61–2.32 |  |
| Highlands | 1.15 | 0.71–1.86 | 1.15 | 0.60–2.18 |  |
| Momase | 0.49 | 0.25–1.02 | 0.61 | 0.28–1.34 |  |
| Islands | Ref |  | Ref |  |  |
| Read newspaper/magazine |  |  |  |  | 0.437 |
| No | Ref |  | Ref |  |  |
| Yes | 1.99 | 1.32–3.01 | 1.27 | 0.69–2.35 |  |
| Listen to the radio |  |  |  |  | 0.349 |
| No | Ref |  | Ref |  |  |
| Yes | 1.61 | 1.05–2.47 | 1.38 | 0.70–2.72 |  |
| Watch television |  |  |  |  | 0.291 |
| No | Ref |  | Ref |  |  |
| Yes | 1.34 | 0.86–2.09 | 0.69 | 0.36–1.36 |  |
| Internet access |  |  |  |  | 0.250 |
| No | Ref |  | Ref |  |  |
| Yes | 2.03 | 1.26–3.25 | 1.41 | 0.79–2.52 |  |
| Own a mobile phone |  |  |  |  | 0.026* |
| No | Ref |  | Ref |  |  |
| Yes | 2.09 | 1.38–3.19 | 1.43 | 1.00–2.55 |  |
| A healthy person can have HIV |  |  |  |  | 0.145 |
| No | Ref |  | Ref |  |  |
| Yes | 1.56 | 0.75–3.22 | 1.95 | 0.79–4.77 |  |
| A person can get HIV by sharing food with an infected person |  |  |  |  | 0.340 |
| No | Ref |  | Ref |  |  |
| Yes | 0.76 | 0.42–1.38 | 0.69 | 0.33–1/47 |  |
| Consistent condom use during sex can reduce HIV risk |  |  |  |  | 0.013* |
| No | Ref |  | Ref |  |  |
| Yes | 1.86 | 1.04–3.33 | 2.18 | 1.18–4.04 |  |
| Age at first sex |  |  |  |  | 0.509 |
| <15 | Ref |  | Ref |  |  |
| >15 | 0.77 | 0.51–1.17 | 0.84 | 0.49–1.43 |  |
| Ever paid for sex |  |  |  |  | 0.036* |
| No | Ref |  | Ref |  |  |
| Yes | 3.56 | 1.85–6.84 | 1.75 | 1.01–5.83 |  |
| Number of sexual partners, including spouse (last 12 months) |  |  |  |  | 0.046* |
| 0–1 | Ref |  | Ref |  |  |
| 2 or more | 1.45 | 0.87–2.40 | 1.37 | 1.00–3.14 |  |
| Consistent condom use during sex |  |  |  |  | 0.050* |
| No | Ref |  | Ref |  |  |
| Yes | 0.69 | 0.45–1.05 | 0.59 | 0.34–1.01 |  |
| Had any STIs (last 12 months) |  |  |  |  | 0.639 |
| No | Ref |  | Ref |  |  |
| Yes | 2.63 | 1.36–5.09 | 0.75 | 0.22–2.54 |  |

\* $p \leq 0.05$
COR, crude odds ratio. AOR, adjusted odds ratio. CI, confidence interval. Ref, reference category.
**Note:** Variables such as ever heard of AIDS and STIs had no responses for “ever tested for HIV” and were excluded in the final analysis.

## Discussion

This study assessed the prevalence of HIV testing and determined important sociodemographic factors, HIV-related knowledge, and sexual risk behaviors associated with HIV testing uptake among young men aged 15–24 years using the PNGDHS data. Findings from this study revealed that only 17.1% of young men in this study had ever been tested for HIV, which is lower than that of research in other countries [37,39,40]. The differences could be attributed to differences in HIV knowledge and awareness and the availability and accessibility of HIV testing services in these countries. There is evidence that young men’s low uptake of HIV testing may be attributed to a lack of access to HIV testing services and a lack of trust in these services [35,41]. In addition, fear of stigmatization, fear of being diagnosed positive for HIV, the perceived risk of sexual exposure, poor attitudes by healthcare professionals, and the perceived psychological burden have been found to impede HIV testing uptake [35]. In PNG, HIV testing services have been more focused on the prevention of parent-to-child transmission [42–44] and key populations (e.g., sex workers) [45,46], while such progress is yet to be achieved with high-risk young populations. Given the importance of early HIV diagnosis and linking access to treatment and care, age-specific policy development is required to increase HIV testing uptake among this priority group.

Consistent with previous studies [35,36,47,48], this study indicated that demographic factors, such as age, marital status, and place of residence, were significant predictors of HIV testing uptake. HIV testing uptake tends to be significantly higher with increasing age, with men aged 20–24 years being 1.18 times more likely to be tested than adolescent men (aged 15–19 years). A plausible explanation could be that men aged 15–19 have a lower self-perceived likelihood of HIV compared to those of increased age, who are more likely to be sexually active and knowledgeable about HIV and its related risks [36,48,49]. It is also possible that they are more likely to have married and have more lifetime exposure to HIV testing services, such as during antenatal care [36,50]. Health education and awareness, including school-based interventions to increase HIV knowledge and risk perceptions, are feasible options that could promote positive sexual risk behaviors and increase HIV testing uptake among young men.

There was a negative association between HIV testing uptake and marital status. The odds of HIV testing remained lower among unmarried young men than those who were married. The result corroborated recent studies in Sub-Saharan Africa [36,51,52]. One possible explanation for this could be that young men frequently encounter challenges accessing available HIV testing services, are dissatisfied with the services provided, and have confidentiality issues [35,48]. Another plausible reason could be that young men have a low-risk perception and are unaware of the physical signs of HIV and its adverse consequences [53]. Evidence has shown that young men with low-risk perceptions who do not have access to HIV testing services are less likely to have received an HIV test in their lifetime [54,55]. Youth-friendly health services can help close the HIV testing gap among young men by addressing access barriers, providing a more accessible, equitable, and effective social environment, and linking to HIV treatment and care.

The results further revealed that the place of residence was negatively associated with HIV testing. Young men who were from rural areas were less likely to be tested for HIV, consistent with previous studies elsewhere [52,56]. The possible reason for this could be due to the availability and accessibility of HIV testing services in urban areas rather than in rural areas, where young men would have fewer opportunities to be tested for HIV. This disparity may also reflect a lack of HIV services, a lack of trained HIV-trained health workers, and inequitable outreach programs to reach young men in rural areas, as previously reported [57,58]. While prioritizing areas that require tailored and youth-specific interventions to improve HIV testing access and uptake, there is a need to further expand HIV services to rural areas. Mobile and rural health outreach programs are critical for reaching young men lacking health care access and increasing HIV testing uptake, and should not be neglected.

In agreement with studies in Kenya and Ethiopia [59,60], the current study found that mobile phone ownership among young men increases the uptake of HIV testing compared to non-mobile owners. Mobile phone interventions in healthcare (mHealth) have shown success in improving access to care and treatment, especially in settings where health disparities are more pronounced [61,62]. Evidence is emerging that the use of mobile technologies remains an effective strategy to reach young people improve access to treatment and retention in care and provide information on HIV services and prevention interventions, including risk reduction [59,63,64]. Furthermore, mobile phones have been shown to address barriers of provider prejudice, stigmatization, discrimination, fear of refusal, lack of privacy, and confidentiality [65,66]. Using mHealth technology provides a potentially promising implementation strategy for interventions to remedy disparities and barriers to HIV testing and the care continuum among young men. The study underscores the significance of preliminary, formative research on population disparities in accessibility, effectiveness, willingness, and use of mobile phones in HIV prevention programs.

In this study, young men who knew that consistent condom use during sex could reduce HIV risk were 2.18 times more likely to be tested for HIV than their counterparts. The results were similar to previous studies [67–69], indicating that having a better knowledge of condom efficacy and effectiveness and sexual modes of HIV transmission were associated with condom use. This study also established that young men who consistently use condoms during sex were less likely to be tested for HIV. This is consistent with evidence suggesting that increased knowledge of HIV and risk perceptions were associated with a positive attitude toward condom use [67,68]. It is also possible that discussion of HIV with a sexual partner, knowing the partner’s HIV status, and being in a steady relationship has been shown to influence the consistent use of condoms [70,71]. Regular health education and information dissemination on condom self-efficacy and risk reduction are crucial for preventing HIV infection among this priority group.

Regarding sexual risk behaviors, young men who had paid for sex were more likely to be tested for HIV than their counterparts. These findings are consistent with previous studies elsewhere [72,73], which suggest that young men who engage in risky sexual behaviors such as sex work and inconsistently use condoms appropriately perceive their risks of HIV acquisition and transmission and, as a result, undertake HIV testing. The fact that they engage in such risky behaviors may imply that their overall risk perception differs from that of those who do not engage in risky behaviors [74]. Strategies for young men who transact sex should not only concentrate on HIV testing and treatment approaches but also on the social contexts in which they engage in transacting sex. Similarly, having multiple sexual partners was significantly associated with HIV testing uptake. This finding corroborates a study in Canada [75] that found an association between HIV testing and having concurrent and multiple sexual partners among men. Having concurrent and multiple sexual partners and risky sexual behaviors are predisposed to HIV and other STIs. Evidence shows that young people who have multiple sex partners and are knowledgeable about HIV and its risks are more likely to utilize HIV testing services [75,76]. The positive association between having multiple partners and the uptake of HIV testing implies that there is a need for developing targeted programs for behavior change and risk reduction for young people in this context.

Contrary to other studies [35,36], no association between the uptake of HIV testing and educational level, employment status, wealth index, mass media exposure, age of sexual debut, or having an STI was found in this study. However, this study has provided useful findings that have the potential to inform the strengthening of current HIV prevention programs to target young men in PNG. Improving HIV testing uptake among young people will play a vital role in reducing the risk of acquiring HIV among young men, hence optimizing their health and well-being [53]. Targeted and coordinated programs aimed at increasing HIV testing services among this priority group remain imperative to achieve the national targets.

### Study strengths and limitations

The study is based on nationwide population data with a nationally representative sample. Also, appropriate estimation adjustments, such as weighting, were employed. However, the study has limitations, and findings should be interpreted cautiously. Given that the data are from a cross-sectional study, it is impossible to prove the temporal relationship. Since the study’s outcome depended on self-reporting, social desirability bias is highly probable, which might have led to under-reporting by the participants. In addition, the dataset did not indicate whether young men were in same-sex relationships (such as men who have sex with men) or not, which could have influenced the uptake of HIV testing.

## Conclusion

Findings from this study showed that one in six young men had ever tested for HIV in PNG, which is well below the national HIV/STI targets. Men aged 20–24 years who owned mobile phones, were aware that consistent condom use reduces HIV risks, had ever paid for sex, and had multiple sexual partners were significant predictors of HIV testing. To increase HIV testing uptake among young men, it is crucial to implement targeted and comprehensive youth-friendly HIV/STI education and sensitization programs and enable more accessible and affordable HIV testing services. Moreover, concerted efforts for outreach and community-based testing programs, especially in rural and prioritized areas where HIV testing services are inaccessible, are feasible options for young people in PNG.

## Supporting information

S1 Table. STROBE Checklist.

S1 Dataset. HIV testing-young men dataset.

## Data Availability

All data produced are available online at: https://dhsprogram.com/

## Acknowledgments

The author would like to thank the DHS program and its partners for the approval to analyze the data. Also, the author is greatly indebted to Dr. Fatch Welcome Kalembo from Curtin University, who provided constructive feedback and suggestions to strengthen the quality of this manuscript.

## Author contributions

Conceptualization: McKenzie Maviso

Data curation: McKenzie Maviso

Formal analysis: McKenzie Maviso

Methodology: McKenzie Maviso

Software: McKenzie Maviso

Validation: McKenzie Maviso

Writing – original draft: McKenzie Maviso

Writing – review & editing: McKenzie Maviso

## Notes

### Competing Interest Statement

The authors have declared no competing interest.

### Funding Statement

This study received no specific grant from any funding agency in the public, commercial, or not-for-profit sectors.

### Author Declarations

The Institutional Review Board of ICF International and the Papua New Guinea Medical Advisory Committee (MRAC) gave ethical approval for this work.

### Summary of Updates

The abstract, introduction, methods, results, discussion and conclusions have been revised to improve the quality of the manuscript.

